# Cytogenetic & Molecular analysis in Premature Ovarian Failure

**DOI:** 10.1101/2021.08.03.21261546

**Authors:** Chetan sahni, Rima dada

## Abstract

**Introduction:** Premature Ovarian Failure (POF) being a heterogeneous genetic disease involves the interaction of multiple genetic defects and environmental factors and has been associated with several chromosomal abnormalities, single gene mutations, and genetic polymorphisms. BMP15 is a member of the transforming growth factor β (TGF-β) family. BMP15 gene product (protein) have 3 domians, mature domain (c-terminal region) of BMP15 binds to receptors located on granulosa cell surface to participate in key steps regarding ovarian function, such as granulosa cell proliferation and follicle maturation, ovulation rate modulation, oocyte competence determination and regulating granulosa cell sensitivity to FSH. Single nucleotide polymorphisms (SNPs) of the BMP-15 gene are associated with POF.

**Materials & Methods:** 30 POF patients and 30 healthy age matched controls were recruited for cytogenetic and molecular analysis. 10 ml whole blood was collected for karyotyping and PCR and PCR was performed for known SNPs of BMP-15 gene (−9C>G, 538G>A, 788insTCT and 852C>T) respectively. Amplified PCR products were sequenced commercially.

**Observation/Result:** Thirty cases (mean age 30 years) and thirty healthy controls (mean age 23 years) were recruited for the study. On cytogenetic analysis 2 cases had a 45, XO chromosomal complement. One case was heterozygous for the SNP (−9C>G) and one control was homozygous for the same SNP.

**Discussion:** The prevalence of this SNP was about 10.7% in cases & 3.3% in healthy controls. This polymorphism in promoter region may cause altered expression of the gene and results in POF.

## INTRODUCTION

Premature ovarian failure (POF) refers to the development of amenorrhea due to cessation of ovarian function before the age of 40 years and is characterized by low estrogen and elevated serum gonadotropin. Elevated levels of Follicle Stimulating hormone (FSH) (≥40 mIU/ml) detected on at least two occasions a few weeks apart, is indicative of premature ovarian failure.^**1**,**2**^ The triad for the diagnosis of POF is amenorrhea, hypergonadotropism, and hypoestrogenism. POF affects approximately 1% of the female population by age of 40 years and 0.1% by age of 30 years worldwide.^**3**^ POF is not the same as natural menopause, as natural menopause is an irreversible condition, while POF is characterized by intermittent ovarian function in one half of young women.^**4**^

In the patients of POF, Hypoestrogenism can be satisfactorily treated by hormone replacement therapy. In contrast, fertility cannot be recovered when the diagnosis of POF (or end-stage POI-premature ovarian insufficiency) is generally reached and is often compromised in the early phases of the disease when the clinical manifestations are absent.^**5**^ The age of onset varies widely, depending on the age of onset, the disorder can manifest as primary amenorrhea, without menarche, or secondary amenorrhea after the pubertal development.^**6**^

Kinch et al., (1965) found that there are two histopathological types of POF, afollicular, and follicular form. In the afollicular form, there is a total depletion of ovarian follicles with permanent loss of ovarian function. In the follicular form, follicular structures are still preserved, hence a possibility of either spontaneous or induced return of ovarian function exists.^**7**^

### Causes of Ovarian Failure

The causes of premature ovarian failure broadly categorized into the genetic and pathological category.^**1**^ Amongst the genetic causes numerical & structural anomalies of the X chromosome adversely affect ovarian function. The most common form of X chromosome defect is Turner’s syndrome. Besides genetic causes, a wide spectrum of pathogenic mechanisms like autoimmune, metabolic (galactosemia), infections (mumps), iatrogenic (anti-cancer treatment) may lead to the development of POF. But, the Genetic causes of POF probably comprise about one-third to one-half of all cases.^**8**^

### Genetic Causes

Chromosomal defects involving the X-chromosome are frequently seen in women with POF. The deletion of X chromosome makes follicle or ovum unstable.^**1**^ In the majority of cases, X chromosome defects cause deletion or disruption of genes located in the critical region of the X chromosome between Xq13 and Xq26, which are critical for ovarian function. Several genes responsible for oogenesis and normal ovarian functions are present on the X chromosome and autosomes.

### Genes Involved

Many genes on the X chromosome and autosomes regulate ovarian function. In a meta-analysis, D. Pu et.al reported that five genes *BMP15, ESR1, FMR1, FSHR*, and *INH* are very important for normal ovarian functions. They also found that single nucleotide polymorphisms and mutations in these genes were found to be significantly more common in patients with POF compared with controls.^**9**^

### *BMP 15* Gene

In our study, we had evaluated the polymorphism in the *BMP15* gene, as the *BMP15* gene has a very important role in ovarian functions. The *BMP15* is a member of the transforming growth factor β (TGF-β) family, which is expressed by oocytes from the early stages of follicular maturation and during all stages of development.10 BMP15 protein can form homodimers (BMP15: BMP15) or heterodimers (BMP15: GDF9). The secreted soluble dimer binds to receptors located on the granulosa cell surface to participate in key steps regarding ovarian function, such as granulosa cell proliferation and follicle maturation, ovulation rate modulation, oocyte competence determination and regulating granulosa cell sensitivity to FSH.^**11**^

In the Sanger sequencing projects of the *BMP15* encoding region in panels of POF patients, more than 15 missense variants were identified.^**12**^ Most of them were located in the protein pro-region. This scenario led to engage the researchers in a recent functional exploration of whether *BMP15* promoter polymorphism might be related to the POF phenotype. It has been also established that this variant modifies the paired-like homeodomain transcription factor 1 (PITX1) binding site and leads to *BMP15* promoter transactivation disturbances.^**13**^ This might suggest that fine-tuning of *BMP15* expression is critical for normal human ovarian physiology.

*BMP15* gene located on the short arm of the X chromosome (Xp11.2) within a ‘POF critical region’.^**14**^ *BMP15* gene has two exons of 328 bp and 851 bp. In humans, mutations in the *BMP15* gene have been found in association with both primary amenorrhea and secondary amenorrhea in several worldwide POF cohorts with a variable prevalence between 1.5 and 12%.^**5**^

In light of these findings, one could hypothesize that *BMP15* variations might play a predisposing role in POF. The functional mechanism by which *BMP15* variants with a proven biological impact may disturb ovarian folliculogenesis is presently unknown. It may be predicted that a diminished *BMP15* paracrine signal in the follicle would involve an impairment of the anti-apoptotic effects on granulosa cells.^**5**^ Indeed, further studies are needed to understand the exact role of *BMP15* variants in POF pathogenesis. Thus in our study, we have evaluated the cytogenetic and molecular basis (mutation in *BMP15*) of disease in cases of premature ovarian failure.

## Materials and Methods

The study population consisted of 30 females of POF with no identifiable etiology after detailed clinical, gynecological evaluation. The control group included 30 healthy females with normal menstrual and fertility history. The relevant information regarding physical examination, clinical investigations, developmental milestones, occupational or environmental exposure to heat, radiation, chemicals, or toxins, were documented in a pre-designed proforma. Written informed consent was taken from each patient and control. The study was approved by the Institute Ethical Committee-A.I.I.M.S. New Delhi (ref-no. IESC/T-150/01.04.2015).

### The Inclusion criteria for study (infertile) group were

only Cases of idiopathic POF with primary and secondary amenorrhea after detailed gynecological & cytogenetic investigations, with no recent history of illness, fever or medication (in past three months).

### The Inclusion criteria for Controls (fertile) group were

Controls who had normal menstrual and fertility history, with no recent history of illness, fever, or medication (in the past three months).

### The Exclusion criteria for the study group and control group

1) Gross dysmorphic abnormalities, structural defects, and systemic infection of the urogenital/reproductive system. 2) History of surgical intervention or trauma of genital/reproductive tract obstruction. 3) History of miscarriage, abortion whether spontaneous or induced. 4) Syndromic cases as Turner and Klinefelter (on cytogenetic evaluation).

For this case-control study, 30 cases of mean age 23 ± 4.15 years were recruited from the Department of Obstetrics & Gynecology, AIIMS, New Delhi, and 30 controls of mean age 25 ± 3.49 years were recruited from a group of healthy wives of cases with male factor infertility & healthy female volunteers from AIIMS, New Delhi. Cases included in this study were presented with premature ovarian failure but on clinical examination, no obvious cause was determined in them and all cases were phenotypically normal. So these cases were found idiopathic and further cytogenetic (karyotyping) and molecular (PCR) analyses were needed in all these cases, While Controls were presented with normal menstrual and fertility history, so cytogenetic (karyotyping) analysis was not done in controls. Three genetic analyses were used to observe the result.

1. Cytogenetic (Karyotyping) analysis in Cases (infertile group)
2. Molecular (PCR) analysis in both Cases (infertile group) & Controls (healthy fertile group)

### 1. Cytogenetic analysis

In all infertile cases, the chromosomal analysis was done to identify any numerical or structural chromosomal aberration. The metaphases were captured under 100X (Olympus optical company, Ltd., Tokyo, Japan). The captured metaphases were analyzed using image analysis software Cytovision 3.7 provided by Applied Imaging (ZEISS microscope, Germany) and classified according to ISCN 2005. At least 50 metaphases were analyzed and karyotyped for each individual.

In karyotyping, it was observed that two cases out of 30 presented with Turner’s karyotype (45, X0). Although, in these two cases Turner’s karyotype (45, X0) was observed but they were phenotypically normal. (fig. 1) These two cases were excluded from the study as cytogenetically normal, idiopathic cases were included in this study. The remaining 28 cases were cytogenetically normal.

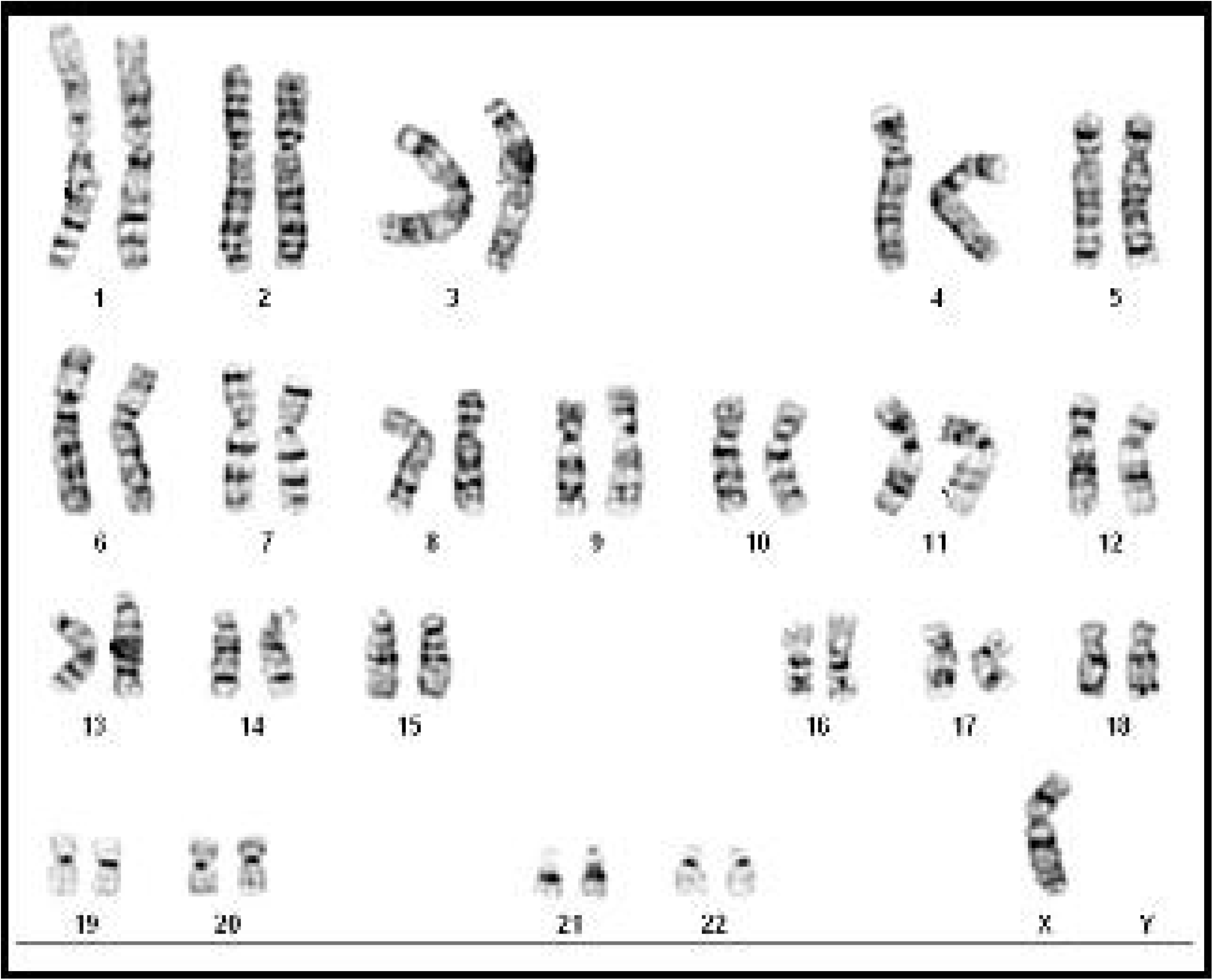

### 2. Molecular (PCR) analysis

SNPs of the *BMP15* gene were observed by using molecular (PCR) analysis. This molecular analysis was done in 28 cases which were found clinically as well as cytogenetically normal but have the phenotype of premature ovarian failure and in 30 healthy fertile controls. Total Genomic DNA was extracted from peripheral blood. Blood samples were collected from cases and controls in EDTA vials only after informed consent. DNA was isolated from 58 (28 cases and 30 controls) samples. Genomic DNA was isolated from peripheral blood samples by miller’s protocol of DNA isolation. DNA was quantified by using a Nanodrop spectrophotometer (Thermo Scientific Corp., USA).

### Analysis of nucleotide variations in the BMP15 gene

The nucleotide variations in the *BMP15* gene were analyzed in cases (n=28) and controls (n=30). In the analysis of the *BMP15* gene, 2 exons (with reference to July 2009 assembly of ensemble.

Amplicons (amplified exons) were electrophoresed using 1.8 % agarose gel. Amplified PCR products were purified using gel/PCR DNA fragments extraction kit (Geneaid Biotech Ltd., Sijhih City Taiwan. Cat no DF100). Purified PCR products were sent for sequencing at MCLAB (Molecular Cloning Laboratories) South San Francisco, CA 94080, U.S.A. (http://www.mclab.com/product.php?productid=19071). DNA sequences were analyzed against BMP15 reference sequence ENST00000252677.3 using ClustalW2 (multiple sequence alignment program for DNA available at http://www.ebi.ac.uk/Tools/clustalw2/index.html provided by European Molecular Biology Laboratory (EMBL) – European Bioinformatics Institute (EBI).

### Statistical Analysis

The data has been expressed as mean ± SD and/or median (minimum, maximum) as per the statistical conventions. Associations were investigated by the chi-square test or fisher’s exact test. SPSS 11.5 (Texas, USA) was used for statistical analysis. Univariate and multivariable logistic regression was done to see the unadjusted and adjusted odds ratio with 95%CI. All the p-values less than 0.05 were taken as significant.

## RESULTS

### MOLECULAR ANALYSIS

By using sequencing analysis software (Chromas pro-2.4), it was observed that, in 3 cases, polymorphism at rs3810682: g.50910775C>G Promoter region of exon -1 of the *BMP15* gene was present. Of the 3 in one case, it was homozygous change while in 2 cases, the heterozygous change was observed. (fig.2)

**Fig. 2:**
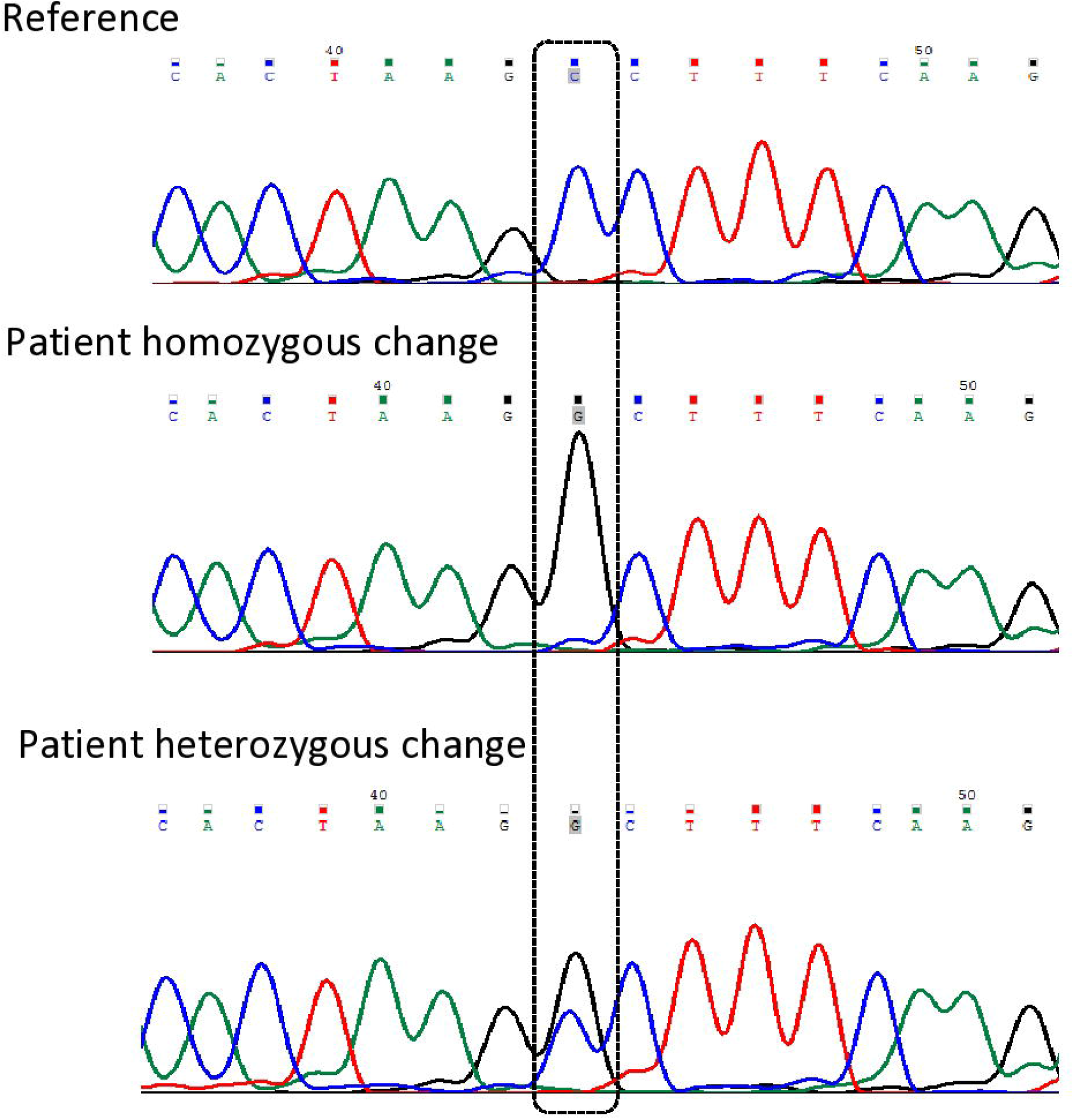
Chromatogram showing sequencing results.

Out of 3 in one case, it was homozygous change. The frequency of the homozygous allele SNPs in cases was 3.7%. In other 2 cases, the heterozygous change was observed. The frequency of the heterozygous allele SNPs in cases was 7%. Besides cases, in one control heterozygous polymorphism was also observed. The frequency of heterozygous allele SNPs in control was 3.3% and frequency of the homozygous allele SNPs in control was 0%. (Table no. 2) All these changes were present at the same location (Promoter region of exon -1 of the *BMP15* gene).

**Table no. 1:**
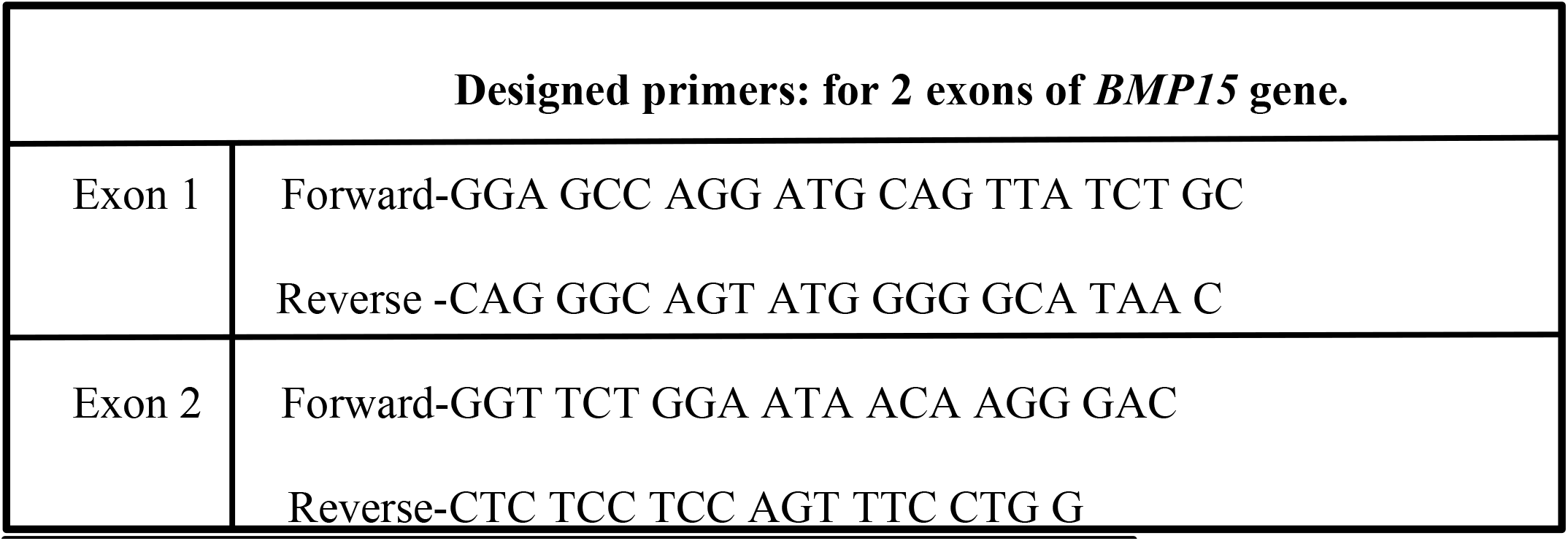
Primers for Exon-1 & Exon-2 of *BMP15* gene.

**Table no. 2:** Allele frequency of SNPs among Cases & Controls.

**Table no. 1: Primers for Exon-1 & Exon-2 of *BMP15* gene**
|  | CASES |  | CONTROL |  | TOTAL |  |
| --- | --- | --- | --- | --- | --- | --- |
| Allele | N | % | N | % | N | % |
| **Heterozygous**<br>**(C/G)** | 2 | 7% | 1 | 3.3% | 3 | 5.2% |
| **Homozygous**<br>**(G/G)** | 1 | 3.7% | 0 | 0% | 1 | 1.7% |
| **Homozygous**<br>**(Reference-Normal)**<br>**(C/C)** | 25 | 89.3% | 29 | 96.3% | 54 | 93.1% |
| **Total** | 28 | % | 30 | 100% | 58 | 100% |

Fisher’s exact test was used to analyze the frequency of SNP variant. It has been found the prevalence of *BMP-15* SNP (rs3810682: g.50910775C>G) among cases was 10.7% and in controls 3.3%. Although, P-value observed 0.344 (at 95% significance level), which was not significant but prevalence was more in cases as compared to controls. (Table no. 3)

**Table no. 3:** *BMP15 GEN*E (Rs3810682: g.50910775C>G) SNPs distribution in cases and controls.

**Table no. 2: Allele frequency of SNPs among Cases & Controls**
| **Group** | ***BMP15 reference gene***<br>(CC-Wild type) | ***BMP15 variant (CG/GG)HOMO/***<br>***HETEROZYGOUS VARIANT*** | **Total** |
| --- | --- | --- | --- |
| Cases | 25(89.3%) | 3(10.7%) | 28 (100%) |
| Controls | 29(96.7%) | 1(3.3%) | 30 (100%) |
| Total | 54(93.1%) | 4(6.9%) | 58(100%) |

## Discussion

The possible involvement of *BMP15* in POF pathogenesis was supported by evidence in animal models.^**15**^ Di pasquale et.al, 2004 reported the first mutation of the *BMP15* gene in two sisters with ovarian dysgenesis. They also described the significant association of heterozygous *BMP15* gene variants with the POF phenotype in humans (seven of 166 patients: 4.2%; *P-*0.003 *vs*. controls). These findings are consistent with the critical role played by *BMP15* in human folliculogenesis.^**16**^

Fonesca et al., 2014, were taken an in-silico approach, which identified that the *PITX1* (pituitary homeobox 1 protein) factor binds to a sequence located between -14 to -8 of the *BMP15* promoter. It was shown that *PITX1* and *BMP15* are co-expressed in both human adult oocytes and in adult mouse ovaries. Functional in-vitro experiments showed that both *BMP15* promoter constructs (*BMP15*-prom-G and *BMP15*-prom-C) were activated by *PITX1*. A statistically significant 1.6 fold increase in *BMP15* transcription activity conferred by the *BMP15*-prom-G construct was found. This feature might lead to alterations in granulosa cell proliferation rate, which may contribute towards POF molecular etiology.^**13**^

In the present study, we, therefore, decided to verify the prevalence of *BMP15* gene alterations among the POF populations. It was observed that both heterozygous and homozygous SNPs (*BMP15 GENE* Promoter region: -9C>G rs3810682: g.50910775C>G) distribution in cases and controls) were present in 3 cases among 28 cases and in one control was observed. The prevalence was higher in cases as compared to controls. By using fisher exact t-test, we observed that although prevalence was higher in cases but p-value was 0.34, which was not significant at 95% CI. This might be due to small sample size. In present study also observed that heterozygous SNPs were more than Homozygous SNPs. Out of 3 in one case, it was homozygous change. The frequency of homozygous allele SNPs in cases was 3.7%. In other 2 cases, the heterozygous change was observed. The frequency of heterozygous allele SNPs in cases was 7%. Besides cases, in one control heterozygous polymorphism was also observed. The frequency of heterozygous allele SNPs in control was 3.3% and frequency of homozygous allele SNPs in control was 0%. All these changes were present at the same location (Promoter region of exon -1 of the *BMP15* gene). Two cases presented with secondary amenorrhea and one with primary amenorrhea. So it suggesting that SNPs associated with *BMP-15* gene cause both primary and secondary amenorrhea.

## CONCLUSION

In previous literature, it has been observed that there is a strong correlation between mutation or SNPs of *BMP-15* gene and premature ovarian failure as the *BMP-15* gene is a regulator of important actions of granulosa cells of the ovary.

In the present study, it was observed that both heterozygous and homozygous SNPs were present in 3 of 28 cases and in one control heterozygous SNPs was observed. The prevalence was higher in cases as compared to controls. By using fisher exact t-test, we observed that although prevalence was higher in cases but p-value was 0.34, which was not significant at 95% CI. This insignificance was due to recruitment of few cases, who presented with idiopathic POF which have normal (46, XX) karyotype. In present study also observed that heterozygous SNPs were more than Homozygous SNPs. Two cases presented with secondary amenorrhea and one with primary amenorrhea. So it suggesting that SNPs associated with *BMP-15* gene cause both primary and secondary amenorrhea Further study, needed with a large cohort of idiopathic cases of premature ovarian failure, to determine the prevalence of allele frequency observed during the study and its genotype-phenotype correlation among both the primary & secondary ovarian failure patients in Indian population and expand mutation spectrum.

## Data Availability

all data available

